# Aerosol Dusters as the Predominant Source of Inhalant Abuse Mortality: Evidence From the U.S. CPSC Clearinghouse, 2011–2021

**DOI:** 10.64898/2026.03.10.26348086

**Authors:** Brian E. Perron, Claudia Dimit

**Author notes:** Correspondence concerning this article should be addressed to Brian E. Perron, School of Social Work, University of Michigan, 1080 South University Avenue, Ann Arbor, MI 48109. **Financial Disclosure** Brian E. Perron has previously served as an expert witness in litigation involving accidents and deaths attributed to difluoroethane inhalant abuse. This manuscript was prepared during a period in which the author was not actively engaged in any such cases. No funding was received for the preparation of this manuscript.

## Abstract

**Background:** Intentional inhalation of 1,1-difluoroethane (DFE), the propellant in aerosol duster products, is a leading cause of inhalant-abuse death in the United States. The CPSC has cited death counts from its Clearinghouse in regulatory proceedings, yet no peer-reviewed publication has described the methods used to identify these cases.

**Objectives:** To estimate DFE- and duster-related deaths in the CPSC Clearinghouse for 2011–2021, characterize reporting patterns, and assess classification reliability against an independently coded dataset.

**Methods:** Death records (N = 6,316) were identified from 261,076 Clearinghouse records using CPSC product codes for chemicals, aerosols, gases, and related products. Each record was classified through narrative review and substance coding. Inter-rater reliability was assessed against an independently coded dataset from Families United Against Inhalant Abuse (FUAIA) using Cohen’s kappa and Gwet’s AC1.

**Results:** Of 2,451 inhalant-abuse deaths identified (70.8% male; mean age 36.9 years), 2,097 (85.6%) involved DFE or aerosol duster products. DFE/duster deaths rose from 110 (2011) to 266 (2016). Only 17% of cases were received in the same calendar year as the incident. Prior to reconciliation, comparison with the FUAIA dataset yielded Cohen’s kappa of 0.90 (95% CI [0.89, 0.91]); all discrepancies were subsequently resolved through joint review.

**Conclusion:** Aerosol duster products account for approximately 86% of inhalant-abuse deaths reported to the CPSC Clearinghouse; however, these counts significantly underestimate true prevalence. The concentration of mortality in a single, widely available product class supports targeted product-level interventions and provides the first peer-reviewed baseline for evaluating the impact of regulatory and prevention efforts.

---

Inhalant abuse, defined as the deliberate inhalation of volatile substances to achieve intoxication, remains a persistent public health concern in the United States. Inhalants are among the most commonly abused substances by adolescents, with the 2022 National Survey on Drug Use and Health estimating that 2.3 million Americans aged 12 and older used inhalants in the past year (1). Unlike most substances of abuse, inhalants can cause sudden death on first use through cardiac arrhythmia. This phenomenon is called sudden sniffing death syndrome (2,3).

Among the diverse substances subject to inhalant abuse, 1,1-difluoroethane (DFE; also designated HFC-152a) has emerged as a substance of particular concern. DFE is the primary propellant in commercially available aerosol duster products (commonly branded as “canned air” or “compressed gas duster”), which are inexpensive, widely available in retail stores, and not subject to age-restricted purchase in most jurisdictions. CPSC staff estimated that approximately 269,000 people per year abused aerosol dusters between 2015 and 2019 (4). From 2012 to 2021, CPSC received reports of over 1,000 deaths and an estimated 21,700 emergency department visits involving DFE inhalation (4). An independent analysis of computer and electronic duster spray-related emergency department visits from the same CPSC data system documented similar patterns (6).

The scale of this problem prompted regulatory action. In April 2021, the nonprofit advocacy organization Families United Against Inhalant Abuse (FUAIA) petitioned the CPSC to ban aerosol duster products containing more than 18 mg of HFC-152a and/or HFC-134a as hazardous substances under the Federal Hazardous Substances Act (5). In July 2024, the CPSC published a proposed rule (89 Fed. Reg. 61363) that would effectuate such a ban, projecting net benefits of nearly $2 billion over 30 years (4). However, in August 2025, the Commission withdrew this and five other pending rulemakings under new leadership, citing a focus on “sound science, robust data, and common sense” and stating that the withdrawn rules “fail to advance safety” (7). The withdrawal was formalized in the Federal Register on September 29, 2025 (8).

Central to this regulatory debate is the death toll attributable to DFE and aerosol duster abuse. The CPSC has reported inhalant death counts derived from its Consumer Product Safety Risk Management System (CPSRMS). This is a surveillance system that includes the Clearinghouse, a national repository of consumer product incident reports compiled from death certificates, medical examiner and coroner reports, and consumer submissions (9). These counts have been cited in briefing packages, rulemaking notices, and the Federal Register. Yet the methodology used to identify inhalant deaths within these data has not been published in the peer-reviewed literature, and the resulting estimates are not part of the scientific record.

This gap matters for several reasons. First, the Clearinghouse was not designed for substance abuse surveillance. Rather, the Clearinghouse is a consumer product safety database with free-text narratives that lack standardized fields for manner of abuse, substance identity, or intent. Identifying inhalant deaths requires reading and interpreting unstructured narratives that use inconsistent terminology across jurisdictions, medical examiners, and reporting years. Second, without published methods, the reliability and reproducibility of reported death counts cannot be independently evaluated, which is a concern reflected in the withdrawal of the proposed rule. Third, there is no published baseline against which future trends or policy effects can be measured.

The present study addresses this gap by reporting the number and characteristics of DFE- and duster-related deaths identified in the CPSC Clearinghouse for 2011–2021, making systematic comparisons across inhalant types, characterizing reporting lag and geographic patterns, and assessing classification reliability by comparing with an independently coded dataset produced by FUAIA.

## Methods

### Data Source

The CPSC National Injury Information Clearinghouse is a federally maintained surveillance system that aggregates consumer product incident reports from multiple sources, including death certificates provided through state health departments, medical examiner and coroner reports, reports submitted via SaferProducts.gov, and news clipping services (9). Each annual dataset contains records from a calendar year (January 1–December 31) with 37 fields, including structured variables (product codes, incident year, victim demographics) and a free-text incident description. The Clearinghouse is a component of the broader CPSRMS. Clearinghouse data for each year generally becomes available in the spring of the following year. All available Clearinghouse records for the period 2011–2021 were obtained, comprising 261,076 total incident records.

### Classification Protocol

Death records were identified from the full Clearinghouse corpus of 261,076 incident records using CPSC product codes associated with chemicals, aerosols, gases, and related products, yielding 6,316 death records. Each death record was then classified through a two-stage process:

1. Narrative review. Each record was reviewed to determine whether it described a death attributable to intentional inhalant abuse. Records were classified as positive (inhalant-abuse-related death) or negative (not inhalant-related or insufficient evidence).
2. Substance coding. Positive cases were assigned a primary substance descriptor: DFE, Duster (product identified as a duster but specific chemical not named), Gases, Gasoline, Nitrous, Paint, Toluene, Other, or Unspecified. For the present analysis, DFE and Duster were combined into a single category representing deaths from aerosol duster products.

### Reporting Lag Analysis

Clearinghouse records contain both an incident date (when the death occurred) and a received date (when the record entered the CPSC system). To characterize the lag between incident occurrence and data availability, cases were tabulated by incident year and received year, and the proportions of cases received in the same calendar year, within one year, and within two years were computed. The estimated completeness of recent incident years was derived from the average lag distribution observed across earlier years (2011–2018).

### Inter-Rater Reliability

FUAIA, a nonprofit advocacy organization focused on preventing inhalant abuse, independently reviewed the same 6,316 death records and classified each as either DFE/duster or other substance, with remaining records classified as negative (5). FUAIA was unaware of the present study during coding, and the two classification systems were developed entirely independently. The comparison was conducted only after both classification efforts were complete. Agreement was computed on the full death-record population (N = 6,316) using Cohen’s kappa (11), Gwet’s AC1 (12), positive agreement, and negative agreement (10). The full death-record population was used as the denominator to avoid the kappa paradox (9,10). After the initial agreement was calculated, all discrepant cases were jointly reviewed, and corrections were applied to the Gold Standard where warranted. The following analyses reflect the corrected Gold Standard.

### Ethical Considerations

This study used de-identified, publicly available administrative data from the CPSC Clearinghouse. No individually identifiable information was accessed. Institutional review board approval was not required. Opus 4.5 (Anthropic) was used for editing the manuscript and assisting with data analysis.

## Results

### Inter-Rater Reliability

The present study’s classifications were compared with the independently coded FUAIA dataset across all 6,316 death records. Prior to any reconciliation, agreement was almost perfect: Cohen’s κ = 0.90 (SE = 0.006, 95% CI [0.89, 0.91]), Gwet’s AC1 = 0.91 (95% CI [0.90, 0.92]), positive agreement = 0.94, and negative agreement = 0.96. Agreement was consistently high across all 11 study years, with annual kappa values ranging from 0.80 to 0.95. All discrepant cases were subsequently resolved through joint review and discussion. The following incident analyses reflect the resolved classifications.

### Inhalant Deaths by Substance Type

From the 6,316 death records, narrative review and discrepancy resolution identified 2,451 inhalant-abuse-related deaths. Table 1 presents the distribution of these deaths by primary substance classification. Of the 2,451 deaths, 2,097 (85.6%) involved DFE or aerosol duster products: 1,216 (49.6%) were coded as DFE (chemical identified in narrative), and 881 (35.9%) were coded as Duster (product identified but specific chemical not named). The remaining 354 cases (14.4%) involved other inhalant substances, including unspecified substances (171; 7.0%), other identified substances (91; 3.7%), gases (41; 1.7%), paint (23; 0.9%), toluene (15; 0.6%), gasoline (11; 0.4%), and nitrous oxide (2; 0.1%).

**Table 1.**
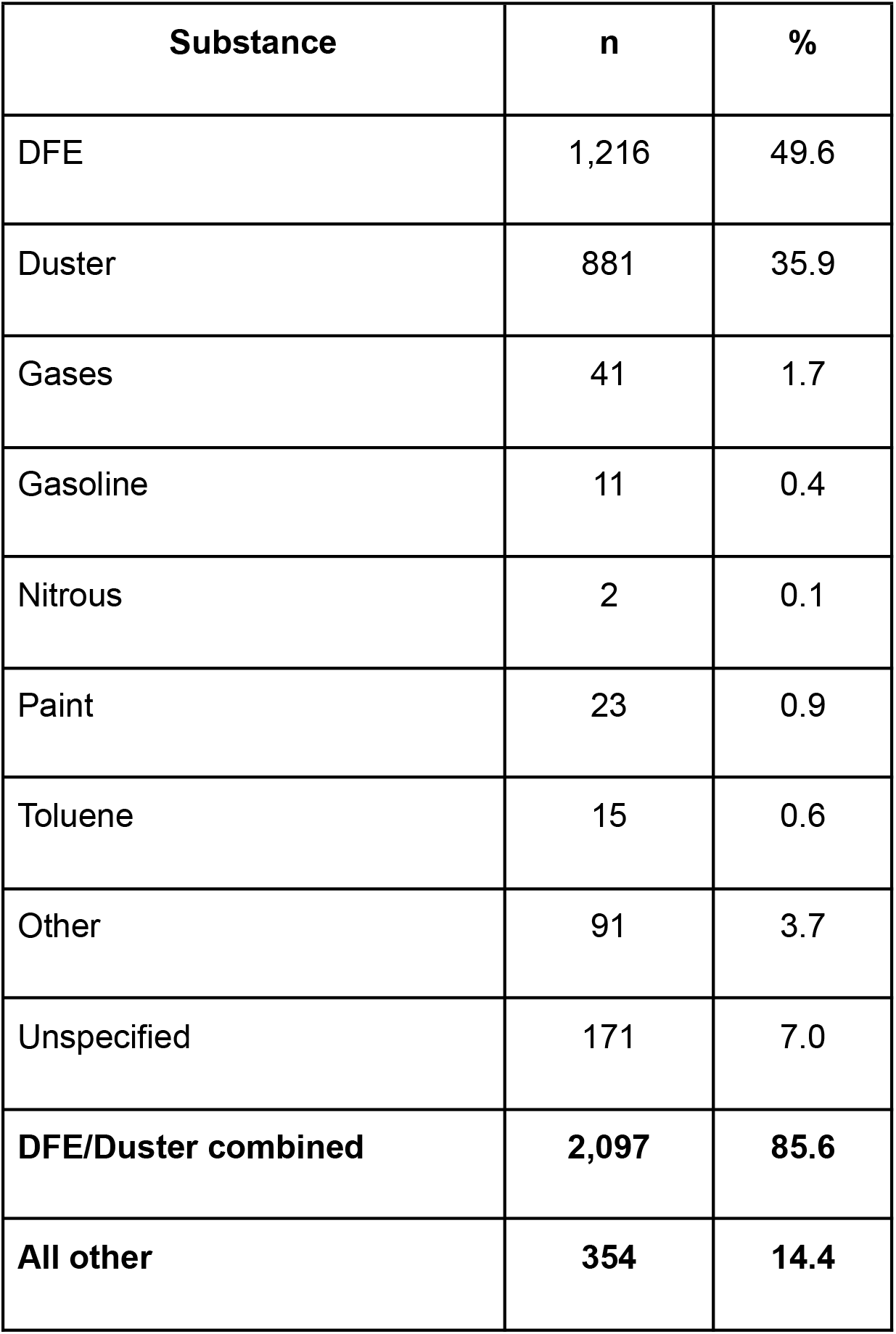
Inhalant-Abuse Deaths by Primary Substance Classification, CPSC Clearinghouse, 2011–2021 (N = 2,451)

### Inhalant Death Trends

Table 2 presents DFE/duster and other inhalant deaths by incident year, which represents when the death occurred rather than when it was reported to the CPSC. After 2017, incident-year counts appear to decline, but this is substantially attributable to reporting lag (see below). The DFE/duster share of all inhalant deaths increased steadily from 74% (2011) to over 90% (2016–2021), indicating the growing dominance of this single product class among reported inhalant fatalities.

**Table 2.** Inhalant-Abuse Deaths by Incident Year and Substance Type, CPSC Clearinghouse, 2011–2021.

| **Incident year** | **DFE/Duster** | **Other**<br>**substance** | **Total** | **DFE/Duster %** | **Est.**<br>**completeness** |
| --- | --- | --- | --- | --- | --- |
| 2011 | 110 | 39 | 149 | 74% | 100% |
| 2012 | 114 | 34 | 148 | 77% | 100% |
| 2013 | 158 | 30 | 188 | 84% | 100% |
| 2014 | 164 | 33 | 197 | 83% | 100% |
| 2015 | 207 | 43 | 250 | 83% | 100% |
| 2016 | 266 | 31 | 297 | 90% | 100% |
| 2017 | 264 | 27 | 291 | 91% | 100% |
| 2018 | 233 | 20 | 253 | 92% | 100% |
| 2019 | 211 | 10 | 221 | 95% | ~94% |
| 2020 | 200 | 12 | 212 | 94% | ~71% |
| 2021 | 54 | 4 | 58 | 93% | ~17% |

### Reporting Lag

The interval between the incident date and CPSC receipt was substantial. On average, only 17% of cases were received in the same calendar year as the incident. The majority of cases (54%) were received in the calendar year following the incident, with an additional 23% received two years later and 6% at three or more years (Table 3). Cumulative receipts reached approximately 71% one year after the incident and 94% two years after.

**Table 3.**
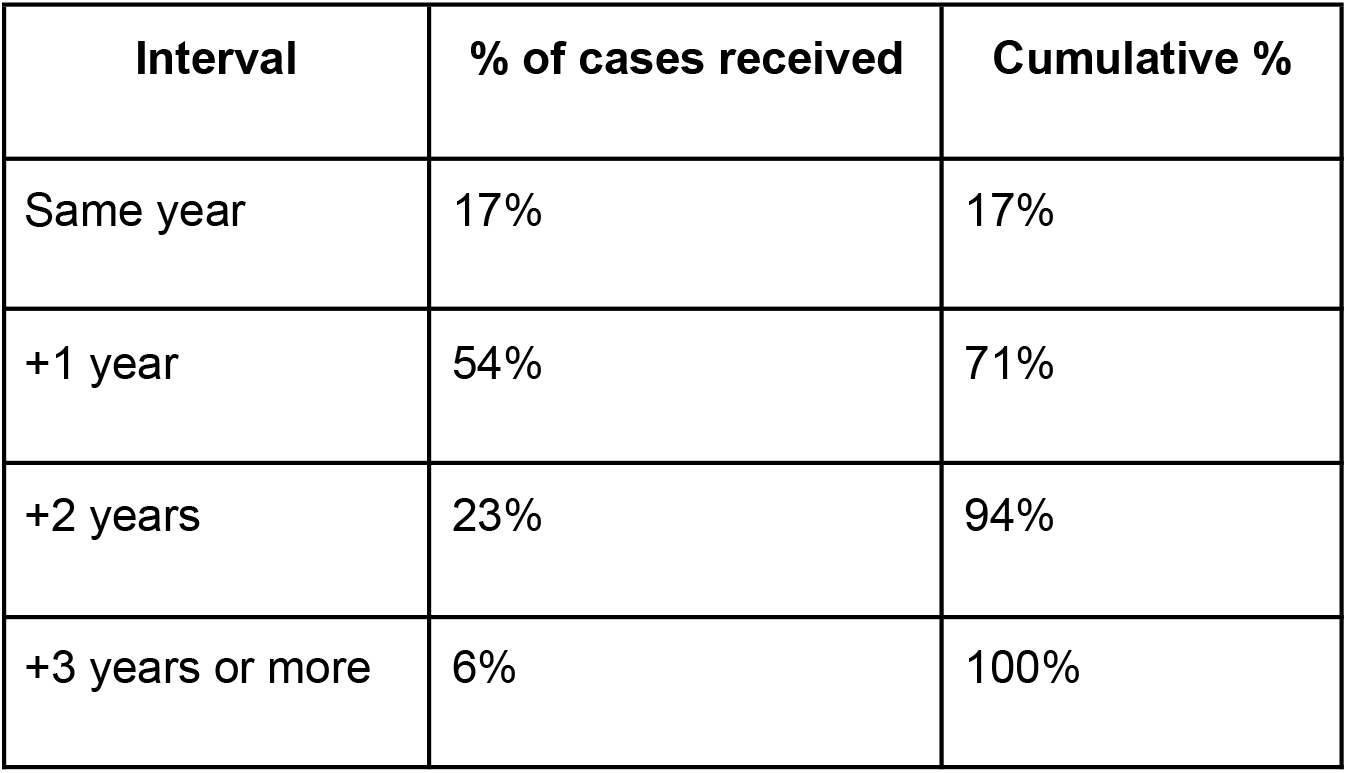
Reporting Lag: Percentage of Cases Received by Interval From Incident Year (Average, 2011–2018 Incident Cohorts)

This lag has critical implications for interpreting recent data. The apparent decline after 2017 is an artifact of incomplete data for recent incident years. The 2021 incident-year count (58 cases) represents approximately 17% of the expected final total — cases received in the same calendar year — because the study period ends with the received year 2021. Applying the average lag distribution, the 2020 incident year is estimated at approximately 71% completeness (∼299 projected cases) and 2019 at approximately 94% completeness (∼235 projected cases). The 2017 and 2018 incident years, with three or more years of follow-up, are essentially complete. Thus, data through 2018 indicate that DFE/duster deaths reported to the Clearinghouse averaged approximately 230–270 per year, with no evidence of decline in the most recent fully reported years.

### Demographics

Among the 2,451 identified cases, 1,736 (70.8%) were male and 710 (29.0%) were female. The mean age at death was 36.9 years (median 36, IQR 29–44, range 8–93). The largest age group was 25–34 years (32.4%), followed by 35–44 (31.3%), 45–54 (18.2%), 18–24 (10.1%), 55+ (6.1%), and under 18 (2.0%).

### Geographic Distribution

Table 4 shows the top reporting states for all inhalant deaths and for DFE/duster deaths specifically. Florida reported the most inhalant deaths by a wide margin (243 total, 223 DFE/duster). These geographic patterns are broadly consistent with prior CPSC analyses of the CPSRMS, which identified Florida, Texas, Illinois, Ohio, Pennsylvania, and North Carolina as the states with the highest counts of aerosol duster reports (4). Minor ranking differences are expected, given differences in time period and classification methodology.

**Table 4.** Top 10 Reporting States, CPSC Clearinghouse Inhalant-Abuse Deaths, 2011–2021.

| **Rank** | **All inhalant deaths** | **N** | **DFE/Duster deaths** | **N** |
| --- | --- | --- | --- | --- |
| 1 | Florida | 243 | Florida | 223 |
| 2 | Texas | 123 | Illinois | 110 |
| 3 | Illinois | 121 | Georgia | 103 |
| 4 | Pennsylvania | 120 | Texas | 94 |
| 5 | Georgia | 114 | Pennsylvania | 92 |
| 6 | California | 104 | Ohio | 87 |
| 7 | Ohio | 100 | North Carolina | 86 |
| 8 | North Carolina | 93 | California | 75 |
| 9 | New York | 70 | Virginia | 66 |
| 10 | Virginia | 69 | Minnesota | 62 |

## Discussion

The present study provides the first peer-reviewed accounting of DFE- and duster-related deaths identified in the CPSC Clearinghouse, covering 2011–2021. Critically, these findings represent a significant underestimate of the true prevalence of inhalant-abuse mortality. The counts reported here reflect only those deaths that were both reported to the CPSC and accompanied by narrative detail sufficient to permit substance classification. Despite this conservative approach, the dominance of a single consumer product class is striking. That aerosol dusters containing difluoroethane account for approximately 86% of identified inhalant-abuse deaths — a proportion that grew steadily over the study period — distinguishes this problem from most substance-abuse domains, where mortality is typically distributed across multiple agents. This concentration has direct implications for how the problem is framed: inhalant-abuse mortality in the United States is, in large part, an aerosol duster problem.

The reporting lag documented in this study introduces a structural limitation that has not been adequately appreciated in prior policy discussions. Regulatory proceedings have treated Clearinghouse counts as if they represent contemporaneous surveillance, yet the data show that an incident year requires at least three years of follow-up before it approaches completeness. Any interpretation of recent trends that does not account for this lag will systematically understate the burden. The CPSC itself has acknowledged the anecdotal nature of CPSRMS data (4), but the magnitude and consistency of the lag have not previously been quantified for inhalant deaths in the published literature. This finding is particularly relevant given the withdrawal of the proposed federal rule, as claims that the problem may be diminishing cannot be evaluated without adjusting for incomplete reporting in recent years.

The geographic concordance between this study and prior CPSC analyses provides an important form of external validation. The two efforts used different time periods, classification approaches, and analytical teams, yet produced closely aligned state rankings. This convergence suggests that the classification methodology employed here is not idiosyncratic but rather captures the same underlying signal that CPSC analysts have identified. At the same time, state-level counts should not be interpreted as incidence rates; the Clearinghouse is an anecdotal reporting system, and cross-state comparisons may reflect differences in reporting infrastructure as much as differences in actual mortality. States with more systematic death certificate submission pipelines to the CPSC may appear to have higher burdens than states with less consistent reporting.

### Independence as Methodological Strength

The inter-rater reliability analysis shows almost perfect agreement between the present study and FUAIA, an advocacy organization that independently classified the same death records using a separate system. Prior to any reconciliation, the agreement was already almost perfect, and all discrepant cases were subsequently resolved through joint review. No communication occurred between the two parties during the coding process; the comparison was conducted only after both classification efforts were complete. This is a stronger form of evidence of reliability than a conventional prospective inter-rater study, in which coders typically share a common training protocol and codebook. Here, a research investigation and an advocacy organization approached the same data from entirely different perspectives and arrived at nearly identical conclusions.

### Translational Implications

The concentration of inhalant-abuse mortality in a single product class has direct implications for prevention and policy. Product-level interventions (e.g., regulatory measures, retail access, labeling) could meaningfully reduce the mortality burden if they specifically reduce misuse of, or restrict access to, duster products. Prevention programs that treat inhalant abuse as a monolithic category may miss the extent to which the problem is driven by one widely available, inexpensive, and unrestricted product type.

The validated methodology also enables future surveillance. With a published, reproducible classification protocol and a demonstrated reliability baseline, subsequent studies can extend the time series, compare counts across data sources (e.g., National Poison Data System, National Vital Statistics System), and evaluate the effects of policy changes — including the withdrawn federal rule and state-level labeling requirements enacted in Minnesota (2024) and Oregon (2025) — on reported deaths. The quantified reporting lag provides a basis for projecting expected completeness of recent data, which is essential for accurate trend interpretation.

## Limitations

These estimates represent a significant underestimate of DFE- and duster-related deaths, and several factors contribute to this undercounting. First, the Clearinghouse captures only incidents reported to the CPSC through death certificates, medical examiner reports, and other submissions; it is not a comprehensive mortality database. Second, a substantial number of Clearinghouse death records contain redacted or partially obscured narrative information.

Many of these redacted records can be reasonably inferred as inhalant-related, particularly given the predominance of DFE and dusters among classified cases; however, they were excluded from the present counts because the available text did not permit definitive classification. This conservative approach prioritized specificity over sensitivity, meaning that the reported figures reflect a floor rather than a ceiling. Third, variability in reporting practices across jurisdictions substantially affects case ascertainment; many narratives reference huffing hydrocarbons or volatile substances without specifying DFE or identifying the product as a duster, even though DFE is the most commonly documented inhalant in these data. Such cases were classified as Unspecified or Other rather than attributed to DFE. Finally, the reporting lag analysis assumes that future years will follow the same lag distribution as 2011–2018, which may not hold if reporting practices change.

## Data Availability

https://www.cpsc.gov/Research--Statistics/Clearinghouse-Online-Query-Tool

## Notes

### Funding Statement

No funding was received for this research.

